# Concurrent comorbidities substantially alter long-term health behaviours and outcomes of headache patients

**DOI:** 10.1101/2020.10.16.20213819

**Authors:** Anthony Nash, Sarah Goodday, Qiang Liu, Alice Fuller, Clare Bankhead, Caleb Webber, Alejo J. Nevado-Holgado, M. Zameel Cader

**Author notes:** Corresponding author and guarantor.

## Abstract

**Objectives:** To investigate factors affecting the long-term health behaviours and outcomes of headache patients and to design a risk-stratification tool.

**Methods:** A population based observational-longitudinal cohort study using primary care electronic healthcare records from the UK Clinical Practice Research Datalink. 550,103 participants with a headache or migraine diagnosis of which 170,336 patients were identified as having recurrent headache-related events. Univariate and multivariable survival analysis was performed to determine factors influencing remission and a neural network classifier was developed.

**Results:** There were almost twice as many female patients (352,330) as males (197,802). The median age at which a patient first saw their GP in males was 38 years, and in females was 37 years. Whilst age, gender and social deprivation significantly influenced the likelihood of seeing a GP, these factors had little effect on likelihood of remission from a period of recurrent headache-related events. In contrast, a comorbidity during the recurrent headache period reduced the risk of remission and increased median survival time from approximately 400-days up 1600. Increasing numbers of comorbidities progressively reduced the hazard risks for remission. The prediction models on remission within two and five years, demonstrated high precision and recall, with an F1 score of 0.795 and 0.887, respectively.

**Interpretation:** Headache patients who suffer comorbidities have a substantively reduced likelihood of remission, highlighting an opportunity to considerably improve health outcomes in recurrent headache patients through addressing multi-morbidity more effectively. The prediction model could be used to help stratify patients most at risk of poor long-term outcomes.

## Introduction

Headache is among the commonest symptoms experienced by people of all ages and gender. For the majority, a headache is self-limiting and a minor inconvenience but, in many people, headaches require analgesic use and/or medical attention. Previous studies have examined types of headache disorders presenting to general practice.^8^ Whilst tension type headache is the most prevalent headache disorder, usually it is a benign condition responsive to simple analgesics. Migraine is the dominant primary headache disorder through a combination of its high prevalence and its debilitating nature.^1^ Migraine disproportionately affects women compared to men, but men with migraine are considered less likely to consult health services or receive appropriate care.^6,7^ 2% of the population experience chronic migraine^2,3^ with a headache for at least 15-days per month.

Furthermore, disability is correlated with increased frequency of headaches.^2^ In a Swiss cohort study of 591 people aged 19‐20, subsequently interviewed across 30-years of follow‐up, 1 in 5 people with migraine followed a chronic course.^4^ Important studies, including the Chronic Migraine Epidemiology and Outcomes and the American Migraine Prevalence and Prevention found more severe headache-related disabilities in those with chronic versus episodic migraine.^5^

People with recurrent headache and migraine frequently attend emergency units and are referred to neurology departments.^11^ Despite consultation rates being relatively high, many patients with migraine symptoms never receive a migraine diagnosis.^10^ The expectation, however, is those with undiagnosed migraine are more likely to suffer ongoing disability and unmet medical need and therefore may continue to present to medical practitioners. The high healthcare use and behaviours of headache patients, results in considerable direct health costs and indirect costs through lost productivity, sick days and absences^1^. Burden and healthcare use of headache patients may differ between subgroups and understanding the differences could improve headache patient management. Comorbid conditions that have been suggested to influence migraine risk, severity or progression to chronic migraine include asthma, ischaemic heart disease, anxiety, depression, arthritis, obesity and non-headache pain conditions.^12–16^ In contrast, hypertension may be inversely correlated with migraine risk.^17^ However, the impact of these comorbidities upon long term health behaviours in headache patients is poorly characterised with no prior studies in primary care populations addressing behaviours and outcomes over several years at sufficient scale. In this study, we used a large real-world dataset to examine how demographic factors and comorbidities affect long-term health behaviours and outcomes of headache patients in a UK primary care cohort.

## Methods

### Data source

UK Clinical Practice Research Datalink (CPRD) is a real-world research service supporting public health and clinical studies. Clinical and referral events are indicated by a *medcode*. Patient records were linked with practice-level Indices of Multiple Deprivation (IMD) scores from 2015, where available.

### Population

Records were retrieved from the February 2018 CPRD GOLD database. A review using headache/migraine codes (provided on request) identified 1,768,360 patients with headache codes and 712,212 patients with migraine codes. We included adults (18-65 years) with their first headache/migraine between 1^st^ January 2000 to 31^st^ December 2016, with at least six months of prior registration and a 12-month follow-up. Older adults were excluded to avoid secondary headaches and the 16-year time frame allowed investigation of long-term health behaviours and headache outcomes. The final cohort selection (550,103) is shown in Figure 1A.

**Figure 1.**
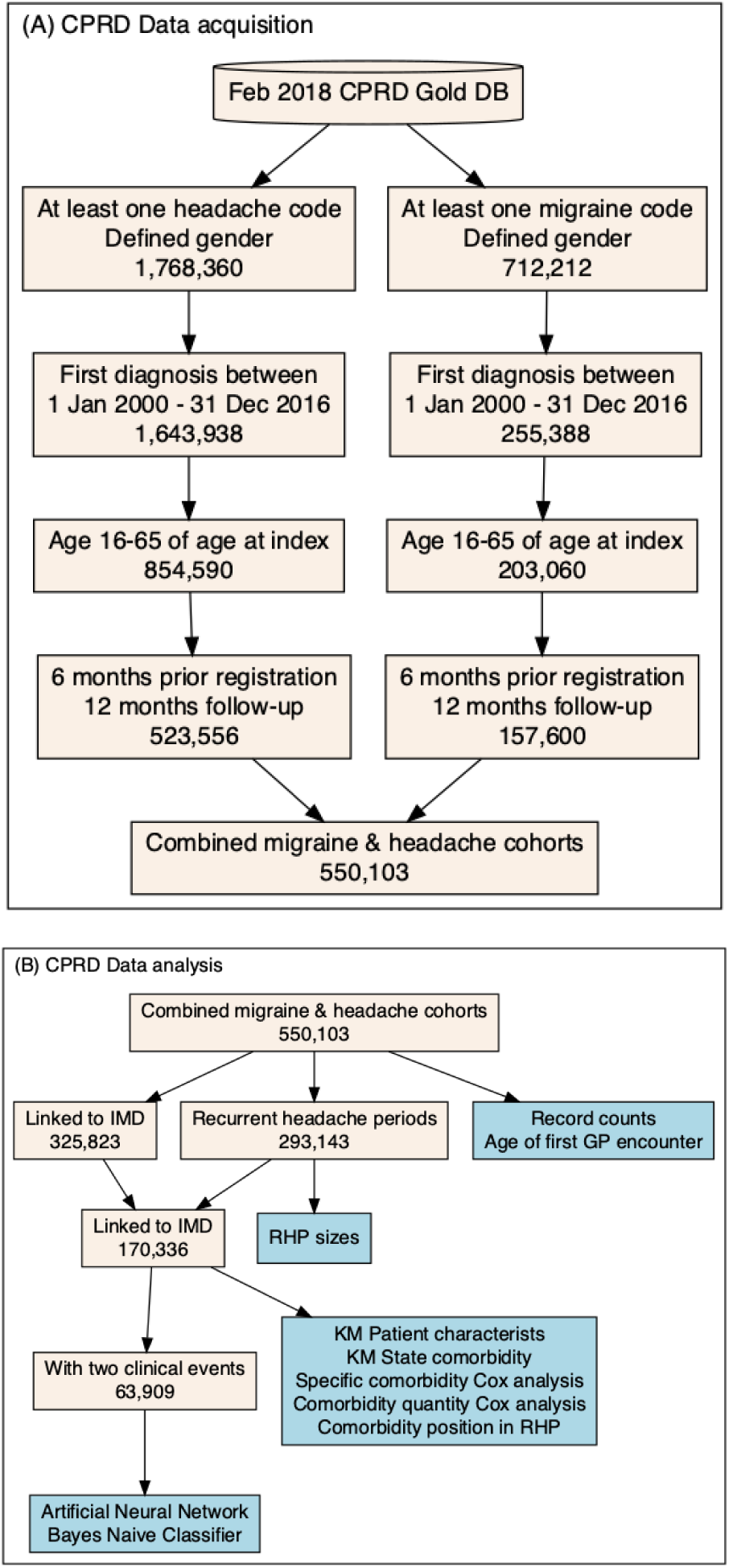
Schematic of (A) the population defined from the CPRD data extraction and (B) the analysis breakdown with corresponding population sizes. 78 patients with neither clinical, referral or therapeutic annotations 240 days after the end of the recurrent headache period, suggesting that these individuals may have died or transferred out of the CPRD, were removed.

We examined three populations (Figure 1B). The first population comprised 550,103 patients. These patient records were linked with IMD scores (325,823). The second population (293,143) comprised a subset of the IMD-linked first population that suffered at least one recurrent headache period (RHP – see below for definition). The third population (170,414) are those who suffered at least one RHP and linked with an IMD score.

### Primary outcomes and recurrent headache period

Medical codes for headache/migraine were identified in patient clinical/referral records. Prescriptions events with product codes for triptans and commonly prescribed analgesics (available on request) were also identified. However, given the almost ubiquitous prescribing of analgesics, these were only considered relevant when the analgesic prescription matched a headache clinical/referral date, and the headache/migraine codes were the only codes recorded on that day (Figure 2 A). Analgesic repeat prescriptions were followed until the prescription came to an end.

**Figure 2.**
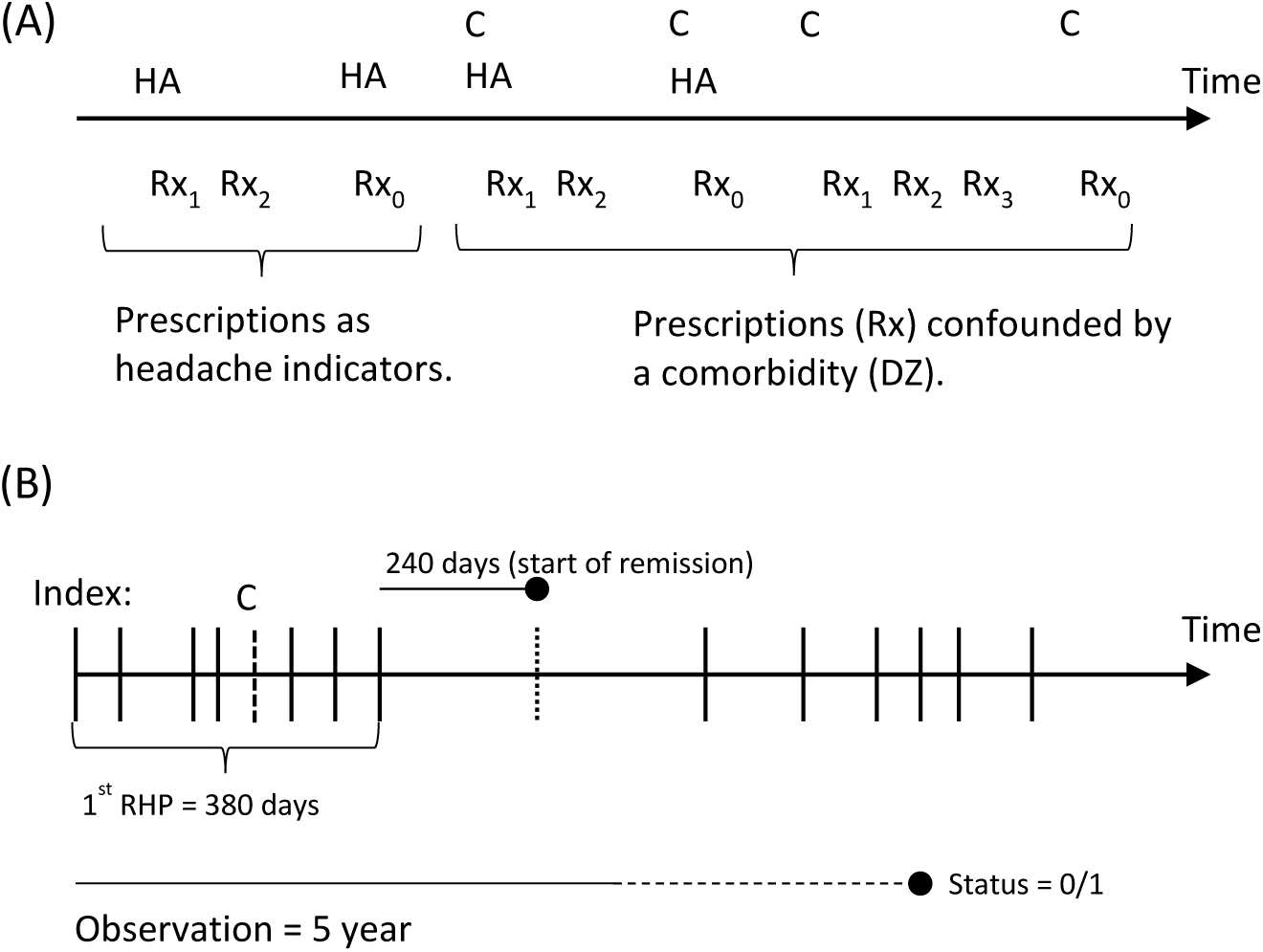
(A) Schematic demonstrating the selection of headache clinical encounters (HA) and analgesic prescriptions (Rx), with a Rx0 indicating a one-off prescription and a Rx1 indicating the start of a repeat prescription. C denotes comorbidities. (B) An illustration of a patient’s record denoting a RHP. The index date is identified as the first headache indicator and the RHP continues until the first break of 240-days between headache indicators, in this example spanning 380-days. An indication of a comorbidity (C) is identified within the RHP. For survival analysis, a status is defined as 1 if the remission occurs within the observation period or 0, otherwise.

We adopted headache clinical/referral events and prescription events, collectively termed as headache indicators, as proxies for whether a patient was continuing to suffer with headaches sufficient to lead to ongoing healthcare use. We selected 240-days to define remission as a reasonable period over which a patient’s headaches may be considered to have resolved or subsided. An RHP was defined as the time-interval starting from index date of first headache indicator until first break of at least 240 days in headache indicators (Figure 2 B). The break of at least 240-days defines a headache remission. During this remission period the patient is at risk of relapse. We identified 293,143 patients with at least one RHP. We only used the first RHP in our survival analysis to avoid confounding issues concerning effects prior consultations and prescription may have on remission outcome.

The primary outcome is the RHP duration, that is, time to remission from index date. The secondary outcomes are age of first GP encounter for a headache, the number of headache related clinical, referral and prescription events over a patient’s CPRD record and the number of RHPs in a patient’s record.

### Covariates and stratifying variables

Covariates include patient age at the start of RHP and at first headache clinical event, and IMD score. For survival analysis we considered a series of comorbidities (Table 2). We only considered comorbidities occurring within a RHP in order to understand the impact of a concurrent diagnosis on outcomes. Patient gender (male and female) and a code for headache or migraine were used as stratifying variables in the Cox proportional hazard regression (Cox PH) multivariable and Kaplan-Meier analysis. Race/ethnicity was not readily available for all subjects and therefore not included.

**Table 2.** Headache, drug and comorbidity indicators in patients with a recurrent headache period. *These patients only have one type of comorbidity during their RHP and are used in the Kaplan-Meier survival curves.

| Patients with a recurrent headache period (linked with IMD), (n=170,336) |  |  |
| --- | --- | --- |
| Male | 55,633 (32.7%) |  |
| Female | 114,703 (67.3%) |  |
| A migraine-only patient | 18,226 (10.7%) |  |
| A headache-only patient | 113,218 (66.5%) |  |
| Triptan | 86,341 (50.7%) |  |
| At least one comorbidity within the RHP | 42,718 (25.1%) |  |
| Comorbidity during RHP | Exclusive* | Total |
| Anxiety | 3,661 (2.1%) | 8,062 (4.7%) |
| Asthma | 1,947 (1.1%) | 4,264 (2.5%) |
| Depression | 6,396 (3.8%) | 12,948 (7.6%) |
| Diabetes (I & II) | 997 (0.6%) | 2,791 (1.6%) |
| Epilepsy | 705 (0.4%) | 1,319 (0.8%) |
| Fibromyalgia | 6,444 (3.9%) | 9,368 (5.5%) |
| Hypertension | 3,980 (2.3%) | 7,453 (4.4%) |
| Neuropathic pain | 1,076 (0.6%) | 2,182 (1.3%) |
| Psoriasis or eczema | 4,116 (2.4%) | 7,564 (4.4%) |
| Severe mental disorder | 195 (0.1%) | 639 (0.4%) |
| Stroke | 266 (0.2%) | 690 (0.4%) |
| TIA | 176 (0.1%) | 497 (0.3%) |

### Statistical analysis

We summarised data as mean or median for continuous variables, and frequencies for categorical variables. The number of patient clinical, referral and therapy events were analysed using a logarithmic transformation. In RHPs, headache indicators were observed over the complete record and number of periods were not adjusted by record length.

The Kaplan-Meier estimator was used to estimate the median probability of survival time to remission. We used the population of 170,336 IMD linked patients with RHPs stratified by gender, by patients with *medcodes* for either headache or migraine, IMD score, and age group. Those same patients were also stratified into non-overlapping comorbidity groups. A reference group was identified from those patients with a RHP and no comorbidity (irrespective of timing). Time was measured in days; 95 % confidence intervals (CI) were based on cumulative hazard and a log-rank test was used to compare curves. Patients numbers per group can be found in the Supplementary Information.

Multivariable Cox PH was used to estimate hazard ratios (HR) and corresponding 95% CI of remission in two models. Both multivariable Cox PH models use comorbidities as time-dependent coefficients and were performed using patients with at least one RHP and with a linked IMD score (170,336 patients). The first Cox PH model uses identified comorbidities (see Covariates and Stratifying Variables) whilst the second model uses a count of comorbidities identified during the RHP. The five-year observation window began from the index date of first headache indicator of the RHP and was used in both models. The Cox PH models were tested for covariate cross correlation and multicollinearity bias (see supplementary Figures 10 to 12 and Tables 3 and 4).

If the RHP came to an end within the observation period a remission event has occurred (status=1), if there is no interruption to the recurrent headache period during the observation period the patient is censored (status=0). were censored. Model covariates include the patient age at start of RHP (continuous) and gender (categorical). The presence of a comorbidity (categorical) was assigned 1 if at least one *medcode* for that comorbidity was found during the RHP whilst inside the observation window. All comorbidities found during the RHP but beyond the observation window were excluded (assigned 0).

Deep artificial neural network (ANN) and Bayes Naïve classifier models were used to predict individual remission status within two and five years. 63,909 patients (at least one RHP and two clinical headache *medcodes* and a linked IMD score) were considered, 67% (42,819) were randomly selected for model training and 33% (21,090) were used for validation.

The ANN models were developed based on the multilayer perceptron (MLP) architecture and contain three hidden layers. Age was considered as numerical inputs. Gender, IMD scores, comorbidities and *medcode* were taken as categorical inputs. Gender, IMD scores and comorbidities were pre-processed using one-hot encoding. *Medcode* was fed into an embedding layer before joining into the first hidden layer. The embedding layers were trained along with the MLP. The two-year remission status was taken as an additional predictor for the five-year remission prediction.Over-sampling technique was applied to train the five-year remission prediction model.

The models generated a probability of remission. The hidden layers rectified linear units as activation functions to bring non-linearity into the models. A Sigmoid activation function was applied to squash the output value. We then plotted receiver operating characteristic (ROC) curves and calculated the area under the ROC curves (AUC) and F1 scores to evaluate our models. All analysis was carried out in TensorFlow and the Bayes Naïve Classifier models used the R mlr package.

### Independent Scientific Advisory Committee

This study was undertaken in preparation and for addressing potential confounds for ISAC project 17_255R2.

## Results

A total of 550,103 patients, aged 18-65 years, had one or more clinical encounters with their general practitioner (GP) for a headache between 2000-2016 (Table 1). There were almost twice as many female patients (352,330) as males (197,802) in keeping with the known gender difference in health-seeking behaviour in headache disorders^18^. From the cohort of 550,103 patients, 293,143 patients had at least two headache events (RHP). Of these, the majority (160,000 patients, 55%) had only one RHP (supplementary information Figure 3) with a median duration of 519-days. 35% of patients with one RHP had a migraine diagnosis (vs 6.2% of patients with only one clinical encounter). 90% of patients with one RHP had a triptan prescription, the migraine specific abortive drug, during the RHP.

**Table 1.** Baseline characteristics. P values calculated using a two-sided Fisher Exact probability test for comparing proportions.

|  |  |  |  |  |  |  |
| --- | --- | --- | --- | --- | --- | --- |
| Patients with clinical events, N=550,103 |  |  |  |  |  |  |
| Male | 197,802 (36.0%) |  |  |  |  |  |
| Female | 352,330 (64.0%) |  |  |  |  |  |
| With a migraine code | 119,074 (21.6%) |  |  |  |  |  |
| With a headache code | 431,029 (78.4%) |  |  |  |  |  |
| Only migraine code in patient record | 58,655 (10.7%) |  |  |  |  |  |
| Only headache code in patient record | 405,625 (73.7%) |  |  |  |  |  |
| Patients with headache and migraine diagnostic codes, N=60,419 (11.0% of patients with clinical events) |  |  |  |  |  |  |
| Migraine on first consult | 27,652 (45.8%) |  |  |  |  |  |
| Headache on first consult | 32,767 (54.2%) |  |  |  |  |  |
| Index of Multiple Deprivation, n=325,823 (59.3%)<br>(IMD 1 – Least deprived. IMD 5 – Most deprived). |  |  |  |  |  |  |
|  | IMD 1<br>N=71,843<br>(22.0%) | IMD 2<br>N=68,632<br>(21.1%) | IMD 3<br>N=67,080<br>(20.6%) | IMD 4<br>N=61,318<br>(18.8%) | IMD 5<br>N=56,950<br>(17.5%) | Total<br>N=325,823 |
| Male | 26,052<br>(36.3%) | 24,785<br>(36.1%) | 23,966<br>(35.7%) | 21,745<br>(35.5%) | 20,228<br>(35.5%) | 116,776<br>(35.8%) |
| Female | 457,91<br>(63.7%) | 438,47<br>(63.9%) | 431,14<br>(64.3%) | 395,73<br>(64.5%) | 367,22<br>(64.5%) | 209,047<br>(64.2%) |
| p value | <0.0001 | 0.0109 | 0.3001 | 0.0008 | 0.0058 | 0.0413 |

**Figure 3.**
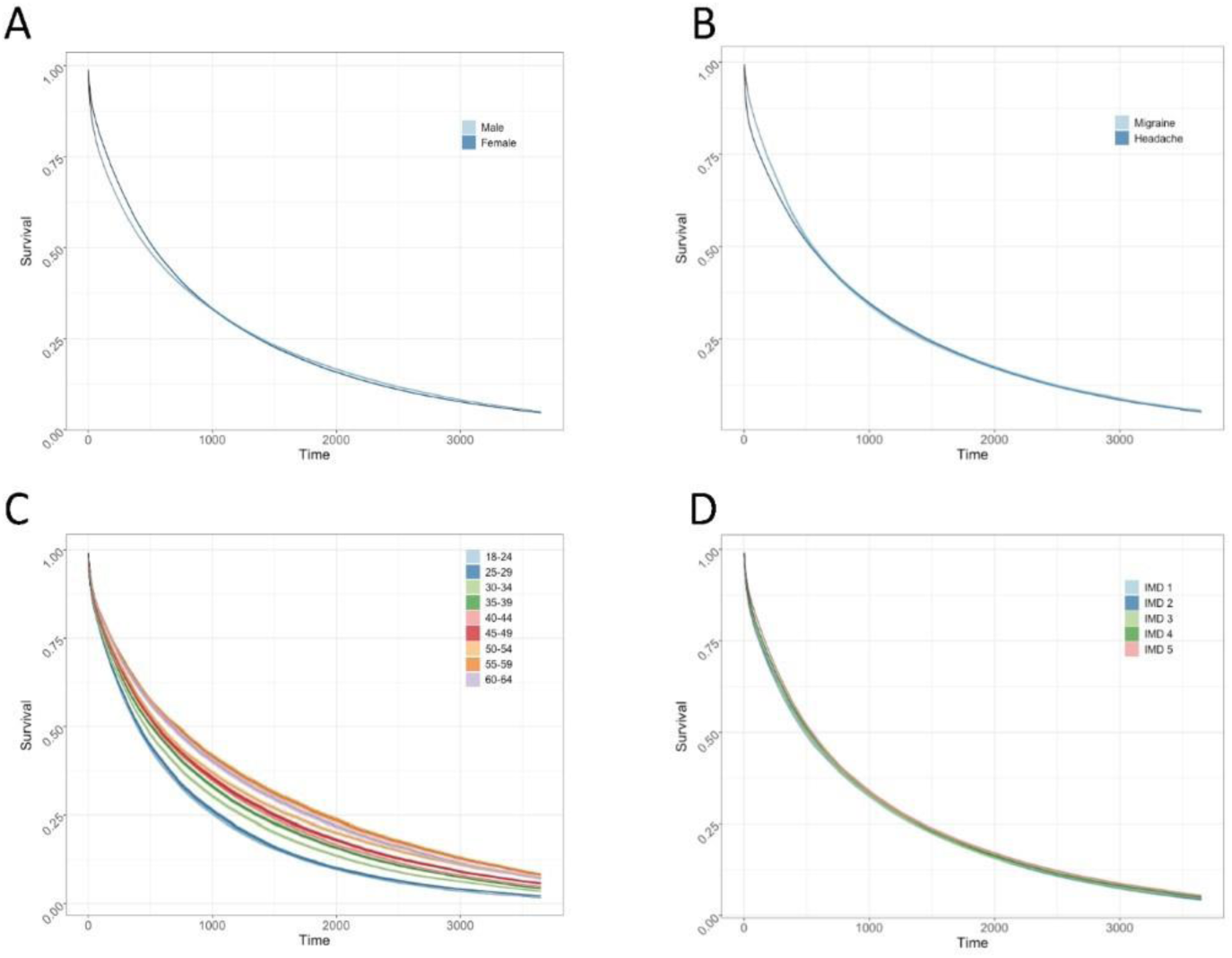
Effect of baseline characteristics on remission, stratified by (A) gender, (B) a history of migraine or headache diagnostic code, (C) the age at the start of the RHP, and (D) IMD score.

The age distribution of first presentation to a GP is also as expected with the majority seeking healthcare between 20-40 years and then steeply declining (supplementary information Figure 1). The median age of first presentation is 38 years for males (first quartile: 30 years and third quartile: 40 years) and is 37 years for females (first quartile: 28 years and third quartile: 48 years).

Most patients were diagnosed with a headache (78.4%) and fewer with migraine (21.6%) and 11.0% had diagnostic codes for both headache and migraine. Patients with a first clinical event of headache were subsequently diagnosed with migraine after a second consultation in similar proportions to those patients initially diagnosed with migraine and subsequently recorded with a headache diagnosis. In the 325, 823 patients where an IMD score as available, there were more patients seeking primary care intervention from the least deprived group (IMD 1 – 71,843 patients) than from the most deprived group (IMD 5 – 56,950 patients).

Most patients only had one clinical encounter, one headache medcode on a referral record and only one prescription for an analgesic (including triptans) (supplementary information Figure 2 A, B and C). There was a significant drop in patients with two or more clinical encounters. 54.8% of patients with one clinical encounter received an analgesic prescription. A clear gender difference was apparent in clinical events, analgesic prescription events and referrals. Women are twice as likely as men to have at least one clinical encounter and receive an analgesic prescription (p<0.0001). The gender difference was apparent across all social groups (Table 1).

In order to understand factors that might influence headache remission for patients in a RHP, we examined the Kaplan-Meier survival times from the first headache clinical encounter to the start of their remission (Figure 3 A–D). We assessed only the first RHP of each patient (170,336 patients, an RHP and a linked IMD score). There were no large differences between survival curves in men and women, although there was a statistically significant difference in median survival time between males (475-days, 95% CI: 465 to 484) and females (527-days, 95% CI: 521 to 532) (Figure 3 A). Interestingly, a migraine diagnosis had no significant impact on median survival time in comparison to patients with a headache diagnostic code (Figure 3 B). IMD score did not significantly affect median survival time. Age of headache onset had a clear influence on median survival time with age group 18–24 years going into remission sooner and age group 55–59 years taking the longest to go into remission.

We next sought to understand the effect of comorbidities on headache remission. Of the 170,336 patients with at least one RHP, 42,718 patients (25.1%) had at least one of the comorbidities we examined. The commonest comorbidity was depression followed by fibromyalgia, affecting 7.6% and 5.5% of the cohort, respectively. There was little collinearity between comorbidities (Supplementary Information for variance inflation factors). The 170,336 patients were divided into subgroups with only one comorbidity or no comorbidities (Table 2). We performed Kaplan-Meier analysis and found that every comorbidity appeared to have a significant and substantial deleterious effect on headache remission (Figure 4). For example, the median survival time for patients with comorbid hypertension was 1603-days (95% CI: 1497, 1705, *P*<.001) and for comorbid depression was 1258 days (95% CI: 1211 to 1315, *P*<.001).

**Figure 4.**
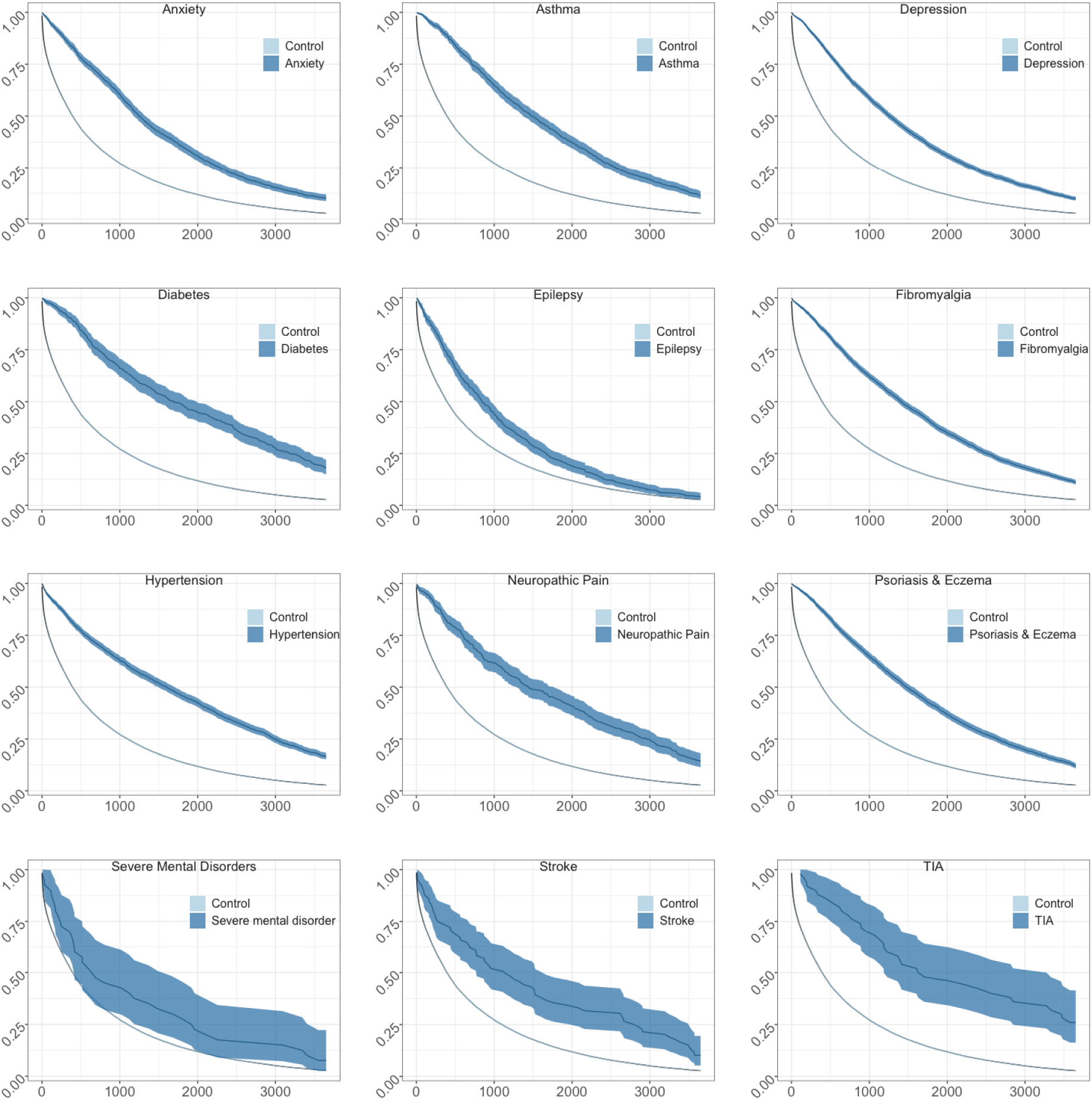
The percentage of patients with non-overlapping state comorbidities by persistent RHPs. The reference group is consistent between measurements and CI (95 %) have been included.

We examined when a comorbidity diagnosis was first recorded within an RHP. The median time after the start of the RHP ranged from 213-days (severe mental illness) to 598-days (TIA). However, in most cases, the comorbidity diagnosis was skewed to the start of the RHP. To further evaluate the time-dependent risk of each comorbidity upon headache remission we used a multivariable Cox PH, including age at start of RHP and comorbidity as covariates and stratified by gender. The effect of gender was small, with Cox PH using a five-year observation window, showing a HR of 0.97 (CI 95%: 0.96 to 0.98, *P*<.001). Comorbidities exerted varying but large effects on risk of remission from the index event. Comorbidities had substantial effects on HR (Figure 5, supplementary Table 1), with psoriasis and eczema (male 1-year and 5-year HR: 0.2360 and 0.6470, *P*<.001, female 1-year and 5-year HR: 0.28 and 0.66, *P*<.001) having the largest impact. Diabetes, asthma, fibromyalgia, hypertension, neuropathic pain and depression also had pronounced effects with HR 0.30 to 0.33 in males and 0.29 to 0.36 in females by one-year. The exceptions were stroke and TIA where HR between males and females diverged substantially.

**Figure 5.**
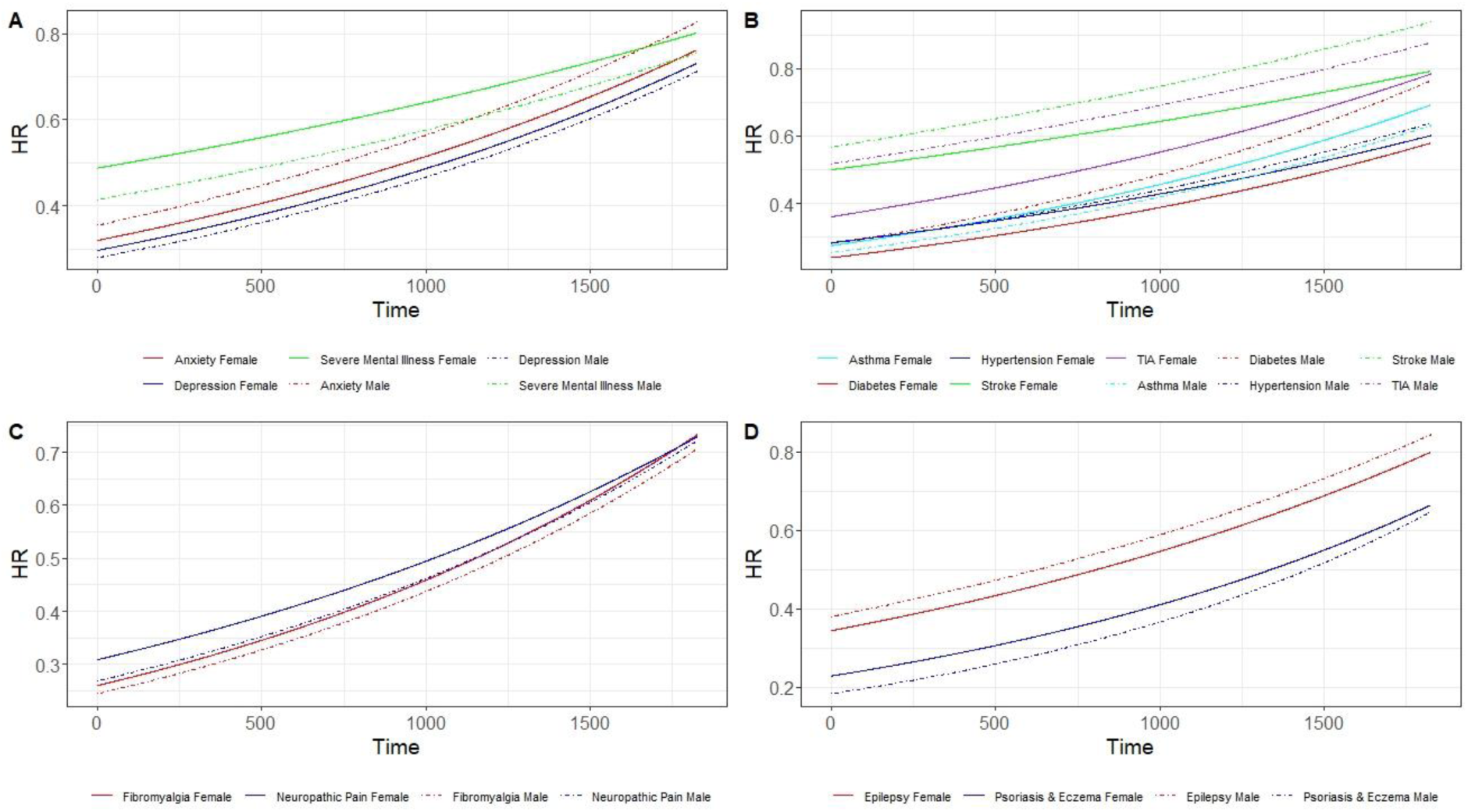
Time-dependent HR of the state comorbidities associated with (A) mental health disorders, (B) vascular disorders and headache risk factors, (C) pain disorders, and (D) other. Cox PH analysis included all comorbidities as covariates in the model but grouped in figure panels for clarity. Each comorbidity was stratified into male (solid-line) and female (dashed-line). Time-dependent HR of the number of comorbidities stratified by (E) male and (F) female.

We next assessed whether multiple comorbidities could further affect remission from a RHP than a single comorbidity. We performed Cox PH using the number of comorbidities as a time-dependent covariate and found the effect on remission became more pronounced (Figure 6, supplementary Table 2); patients with three comorbidities by one-year had a HR less than 0.07 and patients with one comorbidity had a HR less than 0.29.

**Figure 6.**
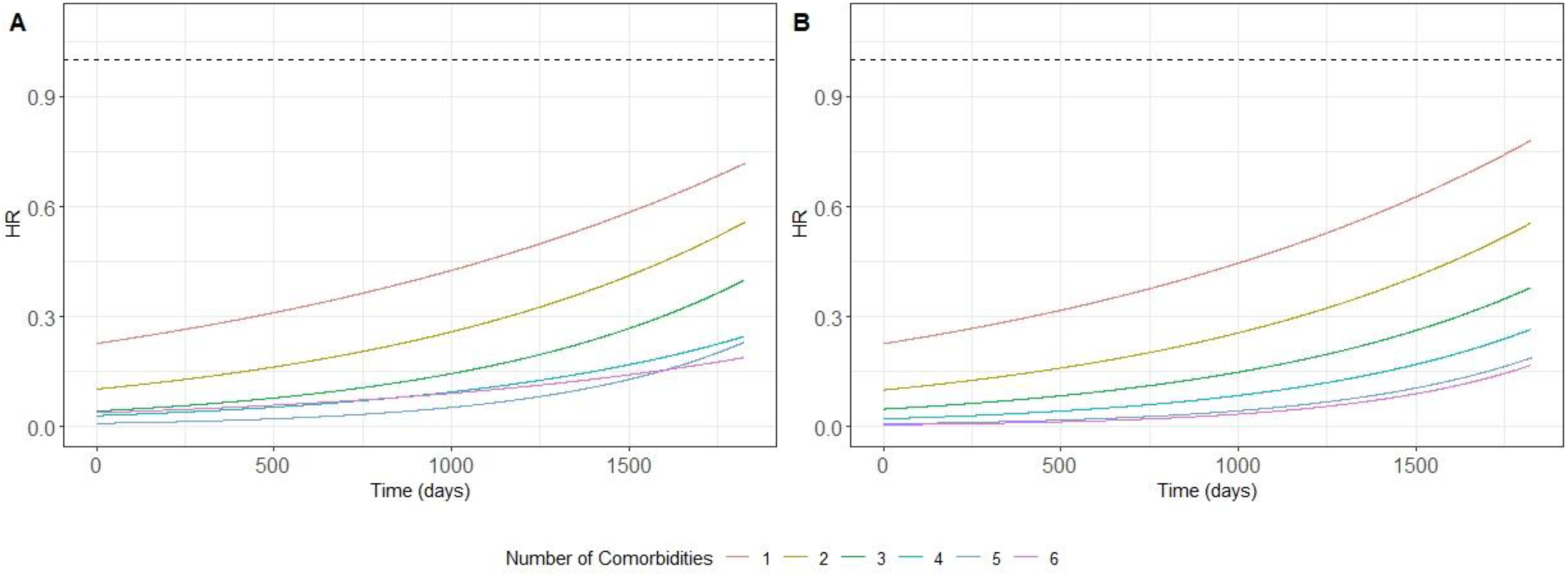
Time-dependent hazard ratios of the number of comorbidities stratified by (A) male and (B) female

Finally, we tested whether we could successfully predict patient remission status within two and five years using an ANN and a Bayes Naïve classifier. We used 21,090 patients who were not involved in model training for testing each method. We then plotted the receiver operating characteristic curves (Figure 7) to evaluate the ANN models. The area under the curve score of the two-year and five-year remission status prediction models are 0.69 and 0.91, respectively. We calculated the F1 score using the cut-off value 0.5 and the results are 0.805 and 0.89, respectively. Detailed confusion matrices and percent change to F1 score by feature are reported in supplementary Figures 13 to 15 and supplementary Tables 5 and 6. The Bayes Naïve classifier returned a-priori distributions of 64.2% and 90.3% of patients going into remission in the two-year and five-year training sets, respectively. The classifier successfully predicted 88.1% and 98.2% of patients going into remission, but only 35.5% and 21.0% failing to remit, over the two-year and five-year test sets (supplementary Table 7).

**Figure 7.**
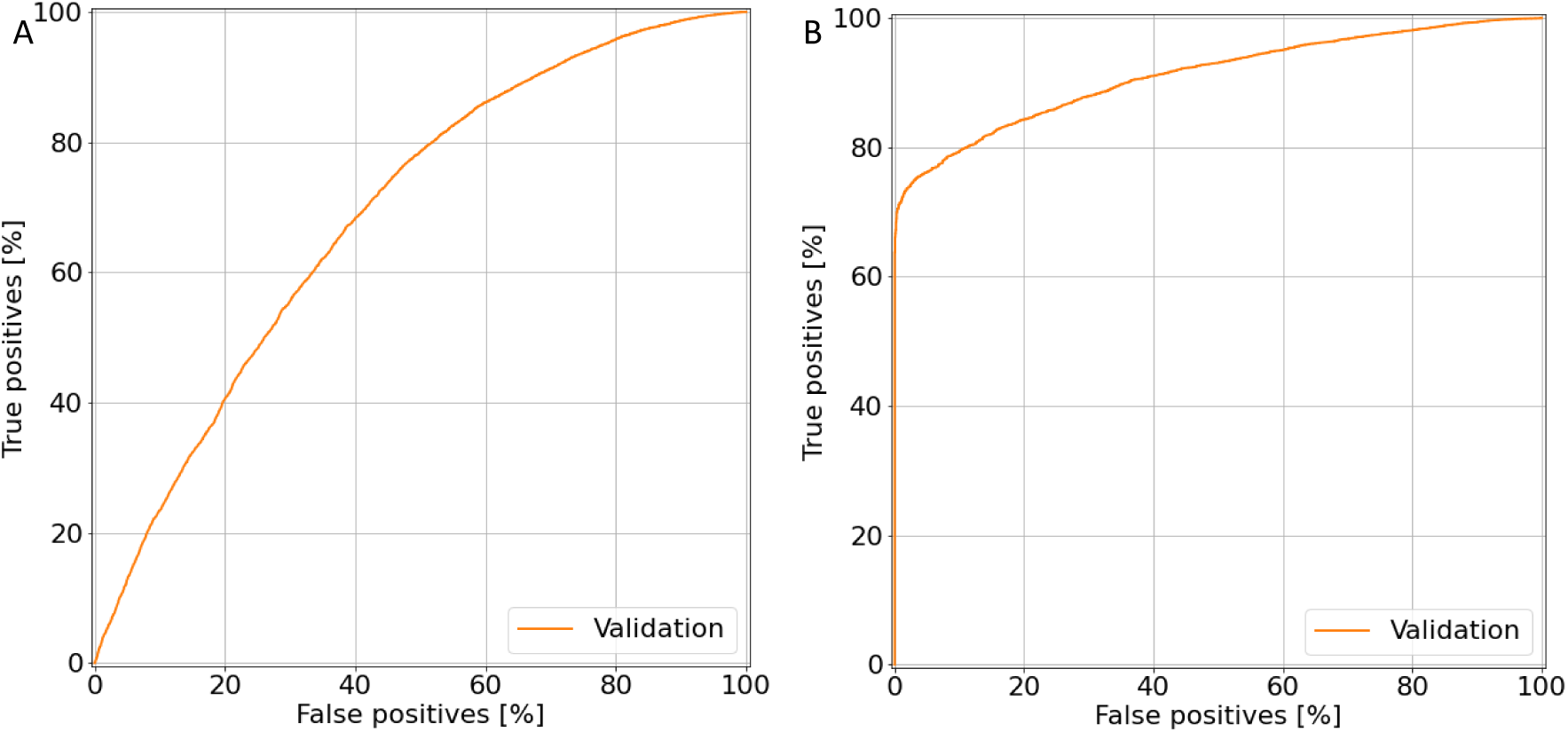
Receiver operating characteristic curves of two-year remission (A) with an area under the curve of .69 and (B) five-year remission with an area under the curve of 0.91 status prediction using the testing set of 21,090 patients.

## Discussion

Prospective studies and population surveys of patients consulting GPs for headache and migraine suggest significant under-diagnosis, suboptimal management and that headache imposes significant disability and impact on quality of life.^19-21^ We examined a large, longitudinal primary care dataset to identify factors influencing headache outcomes. Overall, most patients with headache appear to make minimal use of primary healthcare and likely self-manage with over-the-counter medications or live with headache-associated disability. A coded migraine diagnosis on first presentation is uncommon, but in patients who repeatedly see their GP for headache or obtain prescriptions, a migraine diagnosis increases to 35%. Furthermore, most recurrent headache patients receive a triptan prescription, suggesting that despite a migraine diagnosis frequently not being recorded, GPs are treating ongoing headaches as migraine.

We found the expected gender imbalance in headache prescriptions, with women twice as likely to see their GP and obtain a headache relevant prescription. However, remission times and hazards for remission was not meaningfully different between men and women. This suggests that gender has a significant effect on susceptibility to headache or in health-seeking behaviour but does not affect disease course. Similarly, whilst patients falling into the least deprived category had more clinical encounters, we did not find IMD to affect median remission times. Increased age of first recorded headache diagnosis in contrast increased median survival times in the univariate Kaplan-Meier survival analysis. However, in multivariable models where comorbidity is included as covariates, the magnitude of independent effect of age was small. It is therefore possible median survival times are increased through an increased risk of comorbidity with increased age.

The frequency of comorbidities was low and patients without any comorbidity fared well, with a median survival time of 393-days compared to 500-days in the overall cohort. The effect of a comorbidity on headache outcome was large with median survival time for remission increasing by two to four-fold. In the case of hypertension, the median survival increased from 393-days without any comorbidity to 1608-days in patients developing hypertension during the RHP. The reduced risk of RHP remission was substantial for almost all comorbidities, with the time-dependent Cox PH analysis suggesting that comorbidities independently affect the outcome. We found that as the numbers of comorbidities increased, there was a further decrease in risk of remission. Most likely this is through changes in psycho-social factors leading to changes in health seeking behaviours or prescription habits. For example, headache relevant analgesics may continue to be prescribed simply because other medicines are also being prescribed. Furthermore, the GP may have the patient under increased surveillance.

It is also possible that newly diagnosed comorbidities have a biological effect on headache outcomes through shared genetic factors or through lowering physiological thresholds that trigger recurrent headaches. A shared genetic risk between migraine conditions including hypertension, diabetes, and psychiatric disorders has been demonstrated.^22,23^ The relationship between headache outcome and comorbidity could also be a reverse relationship –individuals with poor headache outcomes are more likely to develop a comorbidity. In this case, recurrent headaches and migraine are a clinical marker of multiple health conditions. It is also possible that recurrent headache disorders, lead to systemic physiological changes that precipitate a liability to other conditions with shared genetic risk such as hypertension or allergic conditions such as eczema.

Based upon the large effects of comorbidity on recurrent headache outcomes, we investigated whether a tool could be developed to predict a poor outcome at two and five years after the first headache clinical encounter. We found the ANN worked well and in future studies should be prospectively evaluated for clinical utility. Although, the Naïve Bayes classifier works well, on the assumption that pairs of features are not independent of each other the model should be considered with care.

The main limitation of our study is that we are unable to directly determine whether a patient is suffering with headache. We only observe health behaviour and it is likely that many patients with headache will be self-treating with over-the-counter medicines. However, the patients we capture are those who feel sufficiently debilitated by their condition that they require health intervention. It is possible also that when patients are in ‘remission’ with respect to their health seeking behaviour, some patients have simply given up due to failed treatments. It is likely that these are patients who then ‘relapse’ and have further RHP as we defined in our study. Also, some stratified comorbidity groups had low samples sizes, likely underpowered as evidenced by wider 95% CI’s and should be interpreted with caution.

## Conclusion

We were able to identify associations between headache related healthcare outcomes and comorbidities. The striking effect of comorbidities during a RHP highlights an opportunity for interventions that could make substantial impacts on the health and disability experienced by headache patients. We were able to develop a stratification tool that could help identify patients at risk of poor outcomes and this merits further evaluation for clinical utility.

## Data Availability

Requests for access to the data reported in this paper will be considered by the corresponding author.

## Acknowledgments

We are grateful to the Oxford Science Innovation, NIHR Oxford Biomedical Research Centre and NIHR Oxford Health Biomedical Research Centre (Informatics and Digital Health theme, grant BRC-1215-20005) for funding. The views expressed are those of the authors and not necessarily those of the UK National Health Service, the NIHR, or the UK Department of Health. Alejo Nevado-Holgado declares support for the work through funding from the European Union’s Horizon 2020 research and innovation programme under grant agreement No. 826421. The funders did not have a role in any part of the study design; in the collection, analysis, and interpretation of data; in the writing of the report; and in the decision to submit the article for publication. The authors would like to thank Dr Michelle Hardy for constructive criticism of the manuscript.

## Contributors

AN was responsible for data acquisition, analysis, design, statistical software development and interpretation and drafting of the manuscript. SG was responsible for analysis and contributions to drafting the manuscript. QL provided neural network analysis and contributions to drafting the manuscript. AJN-H provided statistical contributions. AF and CB were responsible for data acquisition and contributed to writing of the paper. CW was responsible for study conception, design and funding. MZC was responsible for interpretation and drafting of the manuscript, funding, study conception and design.

