## Supplementary material for "Concurrent comorbidities substantially alter long-term health behaviours and outcomes of headache patients": Figures and tables

Tel: + 44 7824499175

#### Contents

1. **Demographic Results**
2. **Kaplan Meier survival analysis**
3. **Time-dependent Cox regression analysis result tables**
4. **Comorbidity Covariate cross correlation and multicollinearity analysis**
5. **Artificial Neural Network and Bayes Naïve Classifier**
6. **References**

### 1. Demographic Results

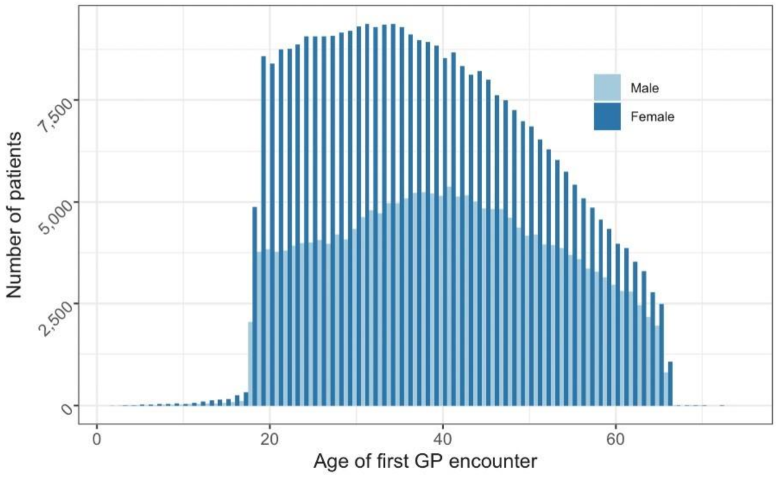

**Figure 1.** Age at first recorded headache or migraine clinical encounter stratified by gender. Age of first GP encounter less than 18 years and greater than 65 years is due to the length of patient record and CPRD selection criteria by age.

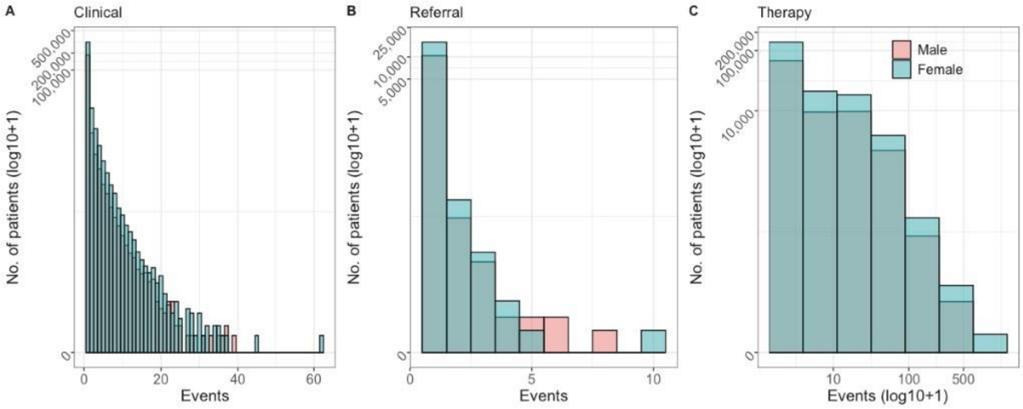

**Figure 2.** The log transformed number of events by gender for clinical events (A), referral events (B) and only therapy events (C).

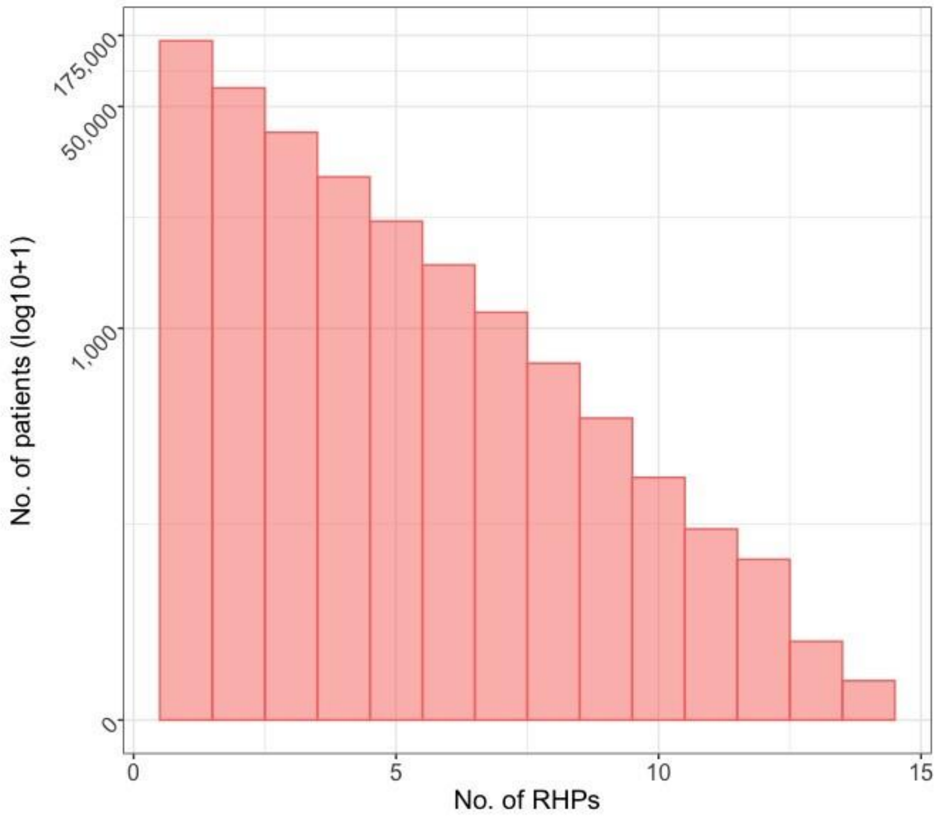

**Figure 3.** The number of patients by the number of RHPs in a patient record.

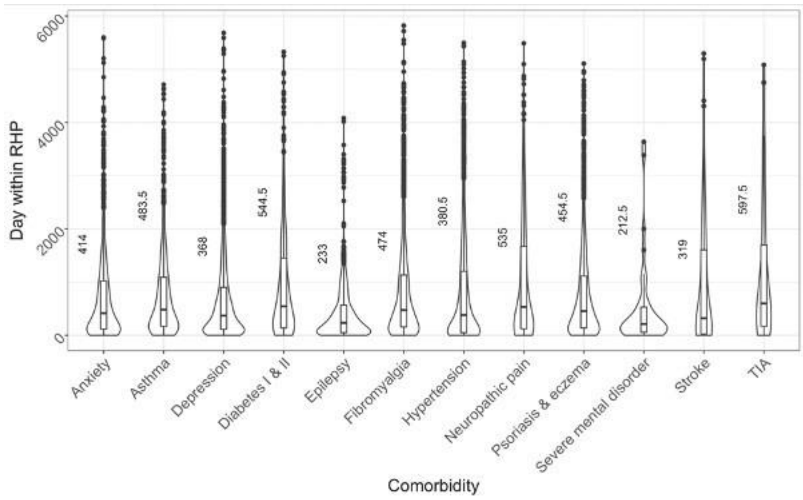

**Figure 4.** The distribution of relative time (in days) from the start of the first RHP to the first instance of each comorbidity. The median values are labelled.-

#### 2. Kaplan Meier survival analysis

##### Number of patients at risk during Kaplan Meier survival analysis

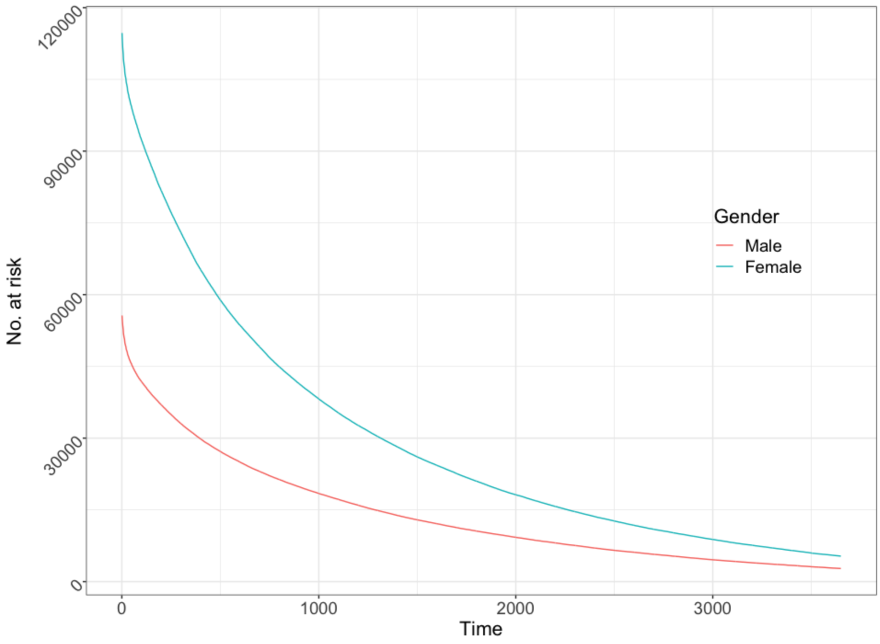

**Figure 5.** The number of patients stratified by gender at risk of a break in survival-time.

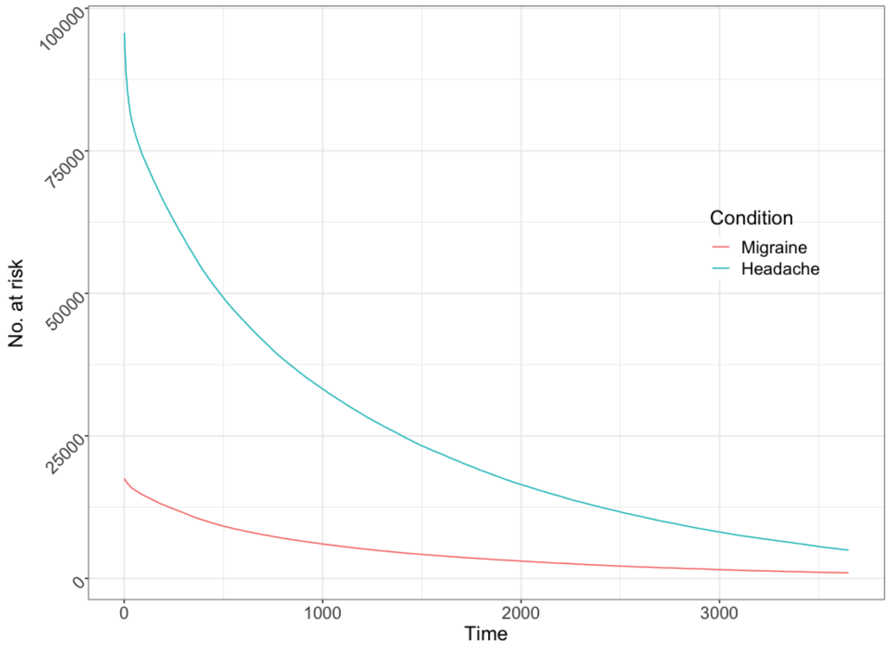

**Figure 6.** The number of patients at risk of a break in survival-time stratified by clinical events which can either be a set of migraine events or a set of headache clinical events, not both.

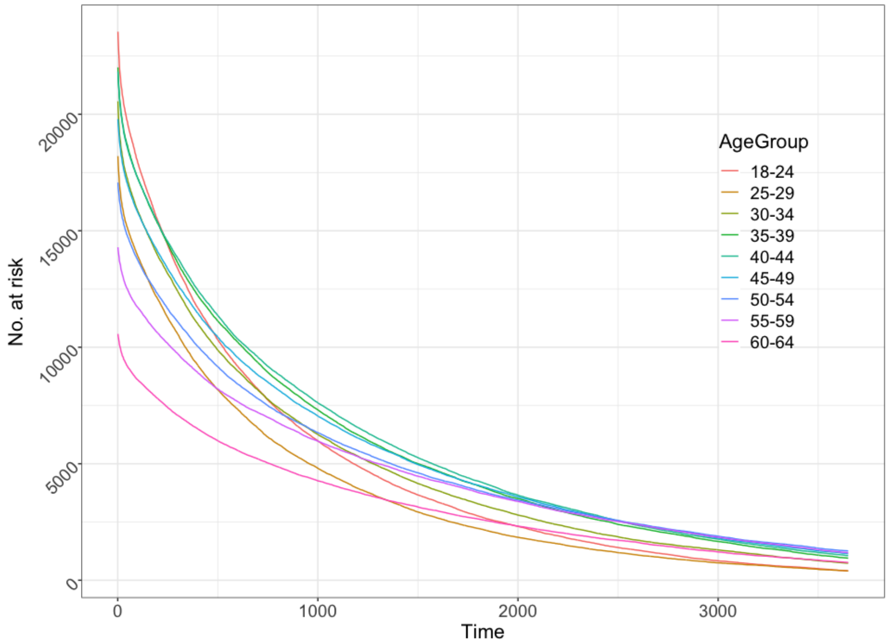

**Figure 7.** The number of patients at risk of a break in survival-time stratified by their age (in years) at the start of a patient's RHP. Once a patient has been assigned an age, they do not move between groups as time increases.

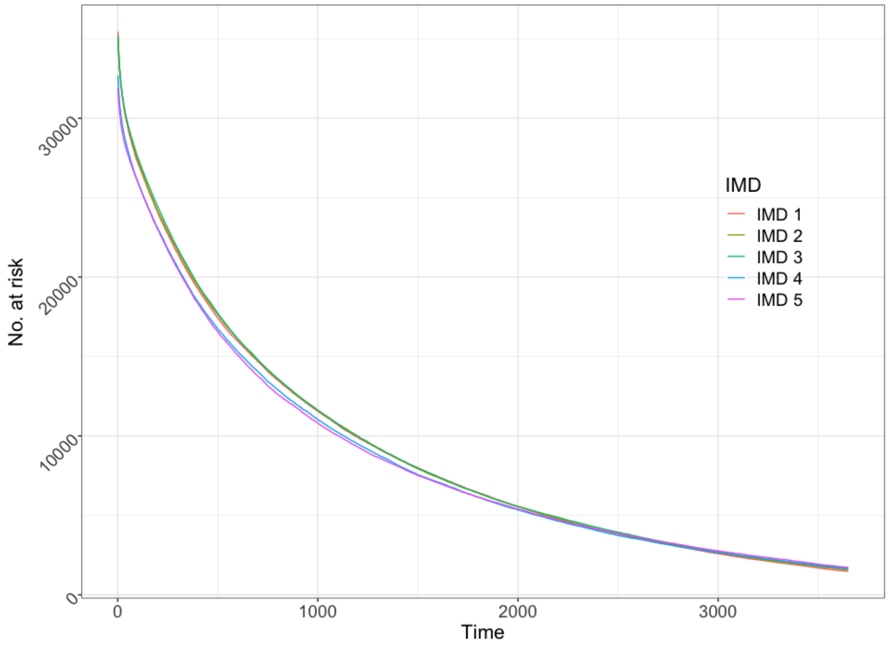

**Figure 8.** The number of patients at risk of a break in survival-time stratified by indices of multiple deprivation.

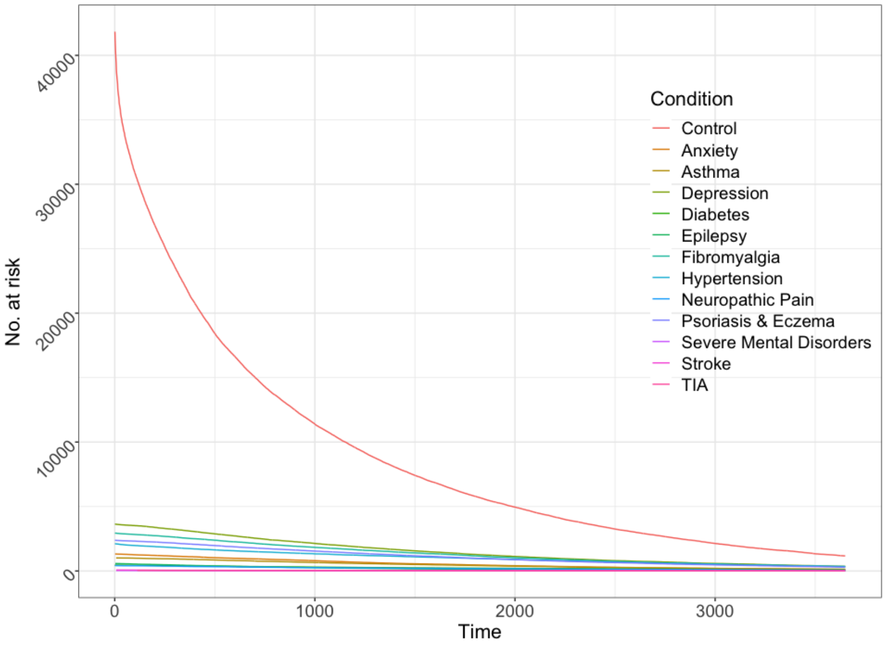

**Figure 9.** The number of patients at risk of a break in survival-time stratified by an indication for a single comorbidity type during their RHP. The control is made of individuals without any indication for comorbidities and patients making up each comorbidity group will only have that one comorbidity during their RHP.

#### 3. Time-dependent Cox regression analysis result tables

Time-dependent HR by comorbidity type.

| | Male | | | | Female | | | |
|---|---|---|---|---|---|---|---|---|
| | **1-year HR** | **2-year HR** | **5-year HR** | **p-value (tt*)** | **1-year HR** | **2-year HR** | **5-year HR** | **p-value (tt*)** |
| **Anxiety** | 0.4209 | 0.4982 | 0.8266 | <0.0001 (<0.0001) | 0.3811 | 0.4533 | 0.7625 | <0.0001 (<0.0001) |
| **Asthma** | 0.3048 | 0.3656 | 0.6311 | <0.0001 (<0.0001) | 0.3308 | 0.3979 | 0.6924 | <0.0001 (<0.0001) |
| **Depression** | 0.3377 | 0.4070 | 0.7120 | <0.0001 (<0.0001) | 0.3560 | 0.4263 | 0.7318 | <0.0001 (<0.0001) |
| **Diabetes** | 0.3425 | 0.4186 | 0.7642 | <0.0001 (<0.0001) | 0.2853 | 0.3406 | 0.5791 | <0.0001 (<0.0001) |
| **Epilepsy** | 0.4453 | 0.5224 | 0.8432 | <0.0001 (<0.0001) | 0.4072 | 0.4820 | 0.7993 | <0.0001 (<0.0001) |
| **Fibromyalgia** | 0.3027 | 0.3742 | 0.7069 | <0.0001 (<0.0001) | 0.3202 | 0.3939 | 0.7333 | <0.0001 (<0.0001) |
| **Hypertension** | 0.3312 | 0.3903 | 0.6384 | <0.0001 (<0.0001) | 0.3296 | 0.3831 | 0.6013 | <0.0001 (<0.0001) |
| **Neuropathic pain** | 0.3277 | 0.3993 | 0.7220 | <0.0001 (<0.0001) | 0.3670 | 0.4858 | 0.7294 | <0.0001 (<0.0001) |
| **Psoriasis & eczema** | 0.2360 | 0.3034 | 0.6470 | <0.0001 (<0.0001) | 0.2829 | 0.3503 | 0.6645 | <0.0001 (<0.0001) |
| **Severe mental illness** | 0.4678 | 0.5273 | 0.7552 | <0.0001 (0.0005) | 0.5389 | 0.5952 | 0.8016 | <0.0001 (0.0001) |
| **Stroke** | 0.6270 | 0.6932 | 0.9373 | <0.0001 (<0.0001) | 0.5479 | 0.6009 | 0.7923 | <0.0001 (0.0006) |
| **TIA** | 0.5749 | 0.6388 | 0.8763 | <0.0006 (<0.0001) | 0.4211 | 0.4920 | 0.7841 | <0.0001 (<0.0001) |

**Table 1.** The HR associated with headache remission in patients with a diagnosis for a comorbidity during the RHP. Discrete HR calculations were taken at one-year, two-year and five-year time-transformations of each comorbidity.

Time-dependent HR by number of comorbidities.

| | Male | | | | Female | | | |
|---|---|---|---|---|---|---|---|---|
| **Number of comorbidities** | **1-year HR** | **2-year HR** | **5-year HR** | **p-value (tt*)** | **1-year HR** | **2-year HR** | **5-year HR** | **p-value (tt*)** |
| **One** | 0.2854 | 0.3594 | 0.7174 | <0.0001 (<0.0001) | 0.2897 | 0.3712 | 0.7812 | <0.0001 (<0.0001) |
| **Two** | 0.1434 | 0.2013 | 0.5568 | <0.0001 (<0.0001) | 0.1407 | 0.1983 | 0.5560 | <0.0001 (<0.0001) |
| **Three** | 0.0661 | 0.1037 | 0.3995 | <0.0001 (<0.0001) | 0.0721 | 0.1093 | 0.3795 | <0.0001 (<0.0001) |
| **Four** | 0.0454 | 0.0692 | 0.2457 | <0.0001 (<0.0001) | 0.0352 | 0.0584 | 0.2657 | <0.0001 (<0.0001) |

| | | | | | | | | |
|---|---|---|---|---|---|---|---|---|
| **Five** | 0.0168 | 0.0323 | 0.2304 | <0.0001 (<0.0001) | 0.0140 | 0.0267 | 0.1876 | <0.0001 (<0.0001) |
| **Six** | 0.0519 | 0.0716 | 0.1883 | <0.0001 (<0.0104) | 0.0100 | 0.0202 | 0.1678 | <0.0001 (=0.0002) |

**Table 2.** The HR for the number of comorbidities identified within a RHP. Discrete HR calculations were taken at one-year, two-year and five-year time transformations of each quantity of comorbidities.

#### 4. Comorbidity Covariate cross correlation and multicollinearity analysis

To determine whether covariates (age, gender, and comorbidities) were subject to multicollinearity we performed three statistical tests over the survival time and covariate values that would later make up the Cox PH regression tests presented in the main text.

##### 1) Comorbidity covariate cross correlation analysis

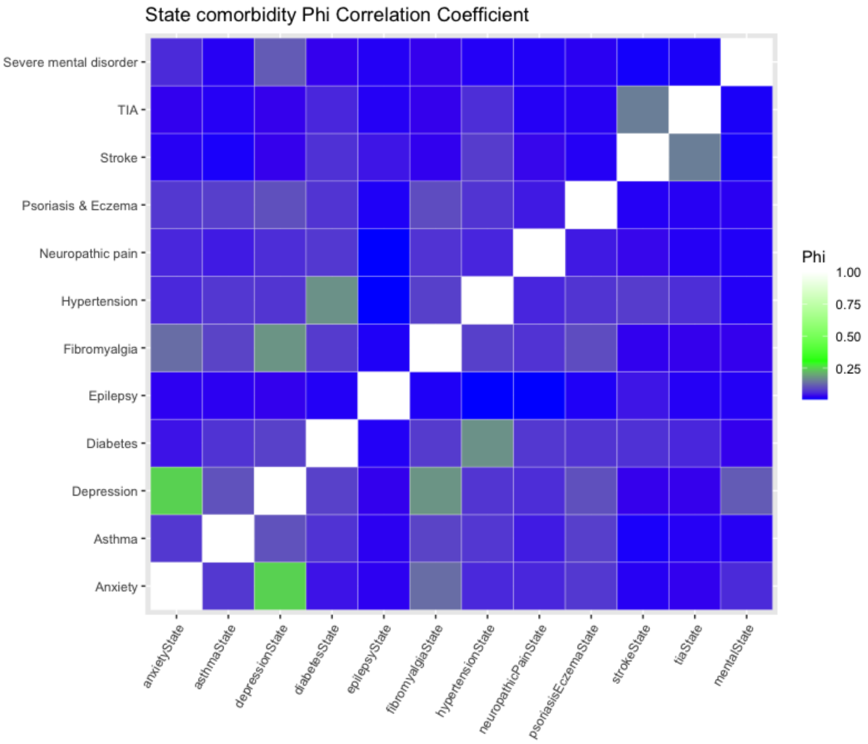

**Figure 10.** Phi correlation coefficient pair-wise analysis for multi-comparison groups between comorbidities identified during the patient RHP survival-time. 1 (white) indicates perfect agreement as per the diagonal identity. In most pair-wise comparisons, Phi ranged from 0 to 0.25, indicating almost no correlation.

**2) Cox model via penalized maximum likelihood:** Glmnet R package.[1,2]

We fit the covariates to a Cox model using the Glmnet R package and visualise the coefficients (Figure 13 and Table 4) followed by performing a cross-validation calculation between covariates (Figure 14).

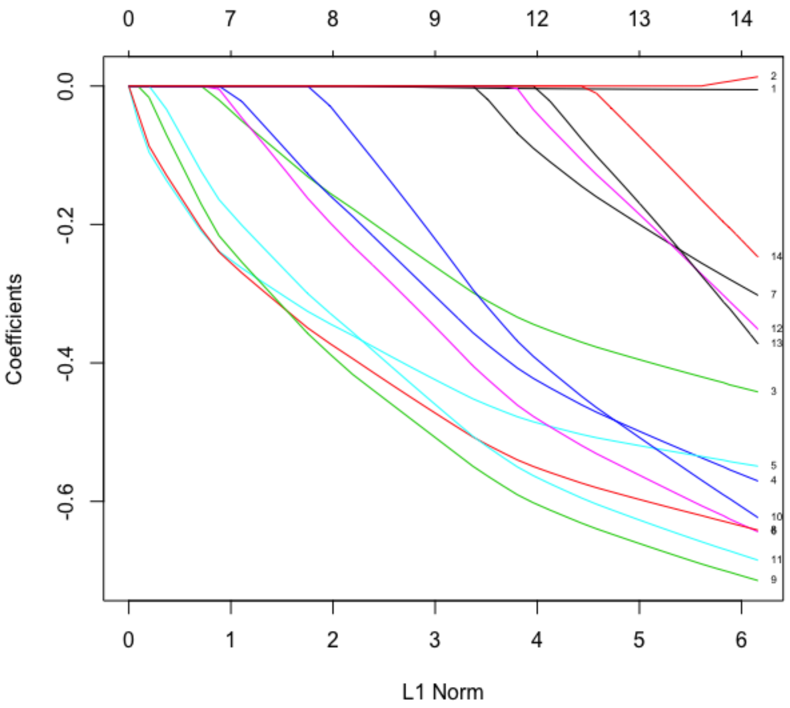

**Figure 11.** Each curve corresponds to one of the covariates; the twelve comorbidity types, age, or gender used in the fitted Cox model.

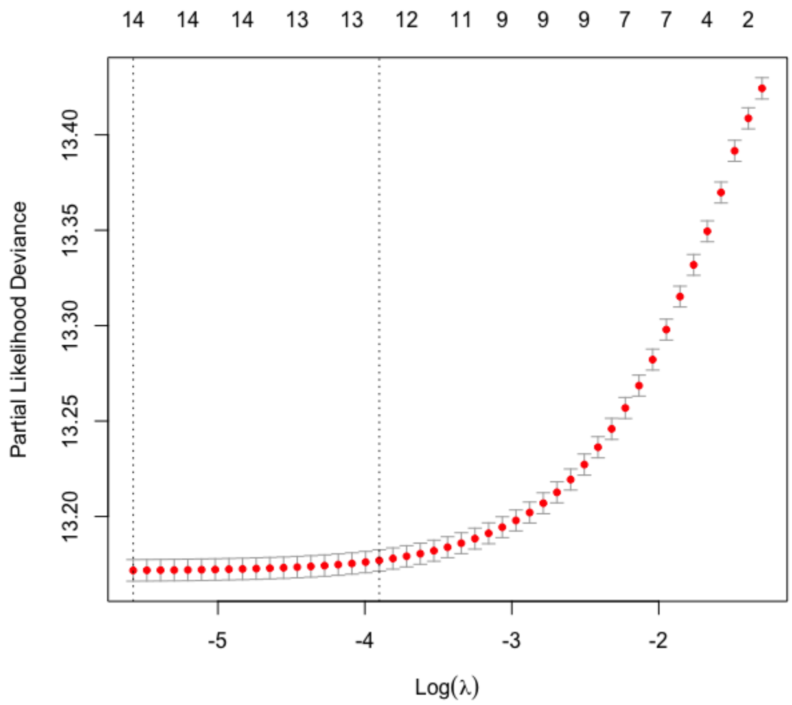

**Figure 12.** The cross-validation curve (red dots), with upper and lower standard deviation along the lambda-sequence. The left-hand dotted vertical line shows where the cross-validation error is at a minimum.

| | **Coefficients** |
|---|---|
| Age | -0.004875828 |
| Gender | 0.019544611 |
| Anxiety | -0.786372949 |
| Asthma | -0.994309957 |
| Depression | -0.895545255 |
| Diabetes | -1.058145995 |
| Epilepsy | -0.532094949 |
| Fibromyalgia | -1.057146063 |
| Hypertension | -0.946808776 |
| Neuropathic pain | -0.959862778 |
| Psoriasis and eczema | -1.134511593 |
| Stroke | -0.453466086 |
| TIA | -0.635270536 |
| Severe mental illness | -0.408558610 |

**Table 3.** The non-zero coefficients for the sampled covariates.

**3) Variance Inflation Factor:** quantifies the severity of the multicollinearity in a regression test using the quotient of the variance in a model with multiple terms by the variance of a model.[3]

| | **Variance** |
|---|---|
| Gender | 1.021330 |
| Age | 1.050546 |

| | |
|---|---|
| Anxiety | 1.033654 |
| Asthma | 1.004862 |
| Depression | 1.050549 |
| Diabetes | 1.021776 |
| Epilepsy | 1.002345 |
| Fibromyalgia | 1.018506 |
| Hypertension | 1.037734 |
| Neuropathic pain | 1.004521 |
| Psoriasis and eczema | 1.003450 |
| Stroke | 1.015059 |
| TIA | 1.012938 |
| Severe mental illness | 1.008774 |

**Table 4.** The variance of each coefficient as a measurement of collinearity. Values less than four are free of collinearity.

#### 5. Artificial Neural Network and Bayes Naïve Classifier

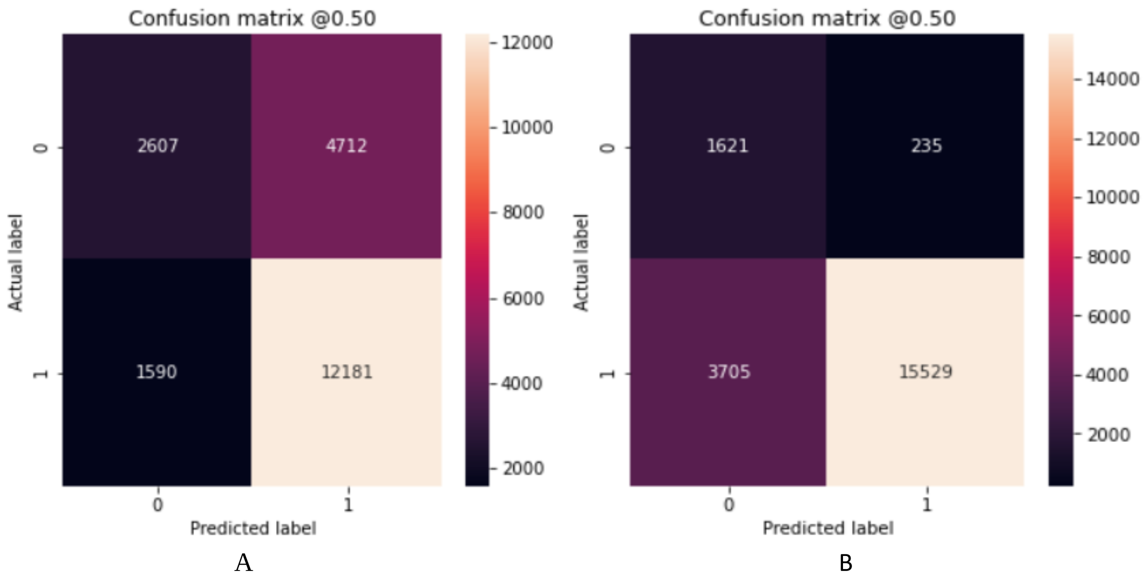

**Figure 13.** Confusion matrices of the internal validation on two-year (A) and five-year (B) remission status prediction models.

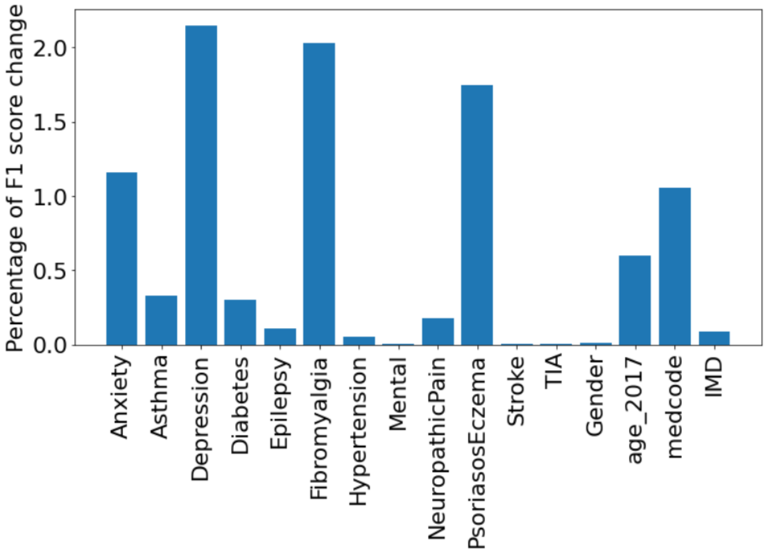

**Figure 14.** Change of F1 score for 2-year remission prediction if each individual input variable is randomly shuffled.

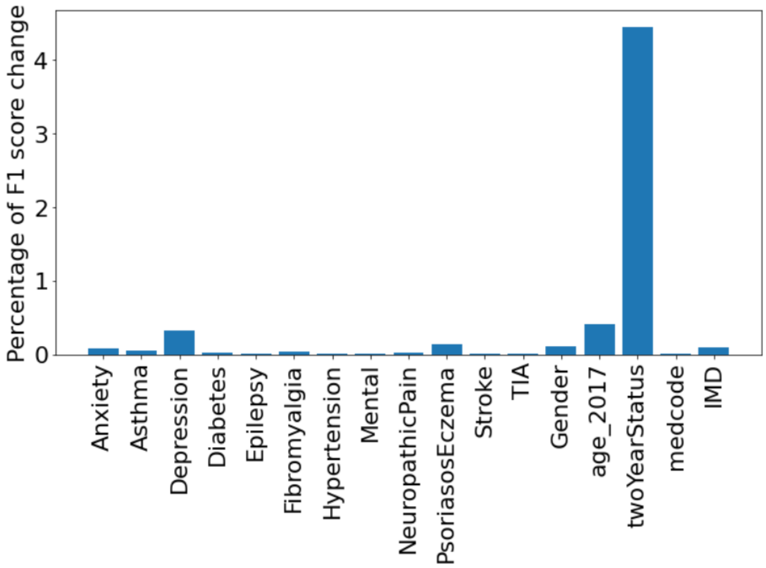

**Figure 15.** Change of F1 score for 5-year remission prediction if each individual input variable is randomly shuffled.

| | Change of F1 score (%) | P-value |
|---|---|---|
| Anxiety | 1.15883 | 3.75E-29 |
| Asthma | 0.329962 | 1.58E-11 |
| Depression | 2.147901 | 3.85E-63 |
| Diabetes | 0.302286 | 9.35E-11 |
| Epilepsy | 0.104485 | 0.001641 |
| Fibromyalgia | 2.03027 | 4.68E-60 |
| Hypertension | 0.054681 | 0.300962 |
| Mental | 0.004852 | 0.712981 |
| NeuropathicPain | 0.173791 | 9.54E-06 |
| PsoriasosEczema | 1.748273 | 6.12E-72 |
| Stroke | 0.001577 | 0.91529 |
| TIA | 0.000757 | 0.910251 |
| Gender | 0.008835 | 0.866698 |
| age_2017 | 0.597057 | 1.15E-08 |
| medcode | 1.05194 | 2.41E-27 |
| IMD | 0.087358 | 0.158088 |

**Table 5.** Change of F1 score for 2-year remission prediction if each individual input variable is randomly shuffled.

| | Change of F1 score (%) | P-value |
|---|---|---|
| Anxiety | 0.08002 | 1.29E-08 |
| Asthma | 0.055703 | 2.35E-05 |
| Depression | 0.319795 | 2.68E-24 |
| Diabetes | 0.022752 | 0.110105366 |
| Epilepsy | 0.004102 | 0.691607964 |
| Fibromyalgia | 0.035039 | 0.0005299 |
| Hypertension | 0.006677 | 0.566987304 |
| Mental | 0.004844 | 0.295342811 |
| NeuropathicPain | 0.015524 | 0.096406972 |
| PsoriasosEczema | 0.135367 | 7.66E-12 |
| Stroke | 0.007807 | 0.108083692 |
| TIA | 0.001946 | 0.668387053 |
| Gender | 0.110552 | 0.010021443 |

| | | |
|---|---|---|
| age_2017 | 0.411983 | 0.000290468 |
| twoYearStatus | 4.446827 | 3.50E-222 |
| medcode | 0.013303 | 0.767162866 |
| IMD | 0.091289 | 0.313252628 |

**Table 6.** Change of F1 score for 5-year remission prediction if each individual input variable is randomly shuffled.

**3) Bayes Naïve Classifier**

| | **2-year** | | **5-year** | |
|---|---|---|---|---|
| **Predictions** | **No** | **Yes** | **No** | **Yes** |
| **No** | 2396 | 1708 | 312 | 359 |
| **Yes** | 4347 | 12640 | 1174 | 19246 |
| **a-priori** | 0.3576 | 0.6424 | 0.0969 | 0.9031 |

**Table 7.** Test set predictions on the two- and five-year model along with a-priori probability distributions of the training-set.
